# Pharmacokinetics of ß-d-N4-hydroxycytidine, the parent nucleoside of prodrug molnupiravir, in non-plasma compartments of patients with SARS-CoV-2 infection

**DOI:** 10.1101/2021.12.06.21267342

**Authors:** Richard FitzGerald, Laura Dickinson, Laura Else, Thomas Fletcher, Colin Hale, Alieu Amara, Lauren Walker, Sujan Dilly Penchala, Rebecca Lyon, Victoria Shaw, William Greenhalf, Katie Bullock, Lara Lavelle-Langham, Helen Reynolds, Wendy Painter, Wayne Holman, Sean Ewings, Gareth Griffiths, Saye Khoo

**Affiliations:** Liverpool University Hospital NHS Foundation Trust, Liverpool, UK; University of Liverpool, Liverpool, UK; Liverpool School of Tropical Medicine, Liverpool, UK; Ridgeback Biotherapeutics, Miami, FL, USA; NIHR Southampton Clinical Trials Unit, University of Southampton, Southampton, UK

## Abstract

ß-d-N4-hydroxycytidine (NHC), the parent nucleoside of molnupiravir, a COVID-19 antiviral, was quantified at sites of SARS-CoV-2 transmission in twelve patients enrolled in AGILE CST-2 (NCT04746183). Saliva, nasal and tear concentrations were 3, 21 and 22% that of plasma. Saliva and nasal NHC concentrations were significantly correlated with plasma (*p*<0.0001).

## INTRODUCTION

An extended therapeutic goal of antiviral therapy is the prevention of infection in individuals that have been exposed to an infected person. SARS-CoV-2 infection occurs through inhalation or inoculation of virus onto upper respiratory airways and mucosal surfaces and in order to be an effective prophylactic agent, drug must penetrate into these sites in sufficient quantities.

Molnupiravir (EIDD-2801; MK-4482), a prodrug of the ribonucleoside analogue 14 β-d-N4-hydroxycytidine (NHC), has recently been licensed in the UK and received FDA Emergency Use Authorisation (EUA) in the US for the treatment of symptomatic COVID-19 in adults with at least one risk factor for developing severe disease. Following oral administration, molnupiravir is rapidly hydrolysed by esterases to NHC, which is phosphorylated by host kinases to active intracellular metabolite EIDD-1931-5’-triphosphate (EIDD-2061) [1]. AGILE, a UK platform for early-phase trials of novel COVID-19 therapies [2], has evaluated molnupiravir within its AGILE Candidate-Specific Trial (CST)-2 seamless phase Ib/IIa protocol. We previously reported phase Ib evaluation of molnupiravir across three dosing arms (300, 600 and 800 mg twice daily), establishing that 800 mg twice daily for 5 days was suitable for progression to phase II [3], which is currently recruiting.

We report here the pharmacokinetics of molnupiravir and NHC in saliva, nasal secretions and tears, in comparison with plasma concentrations within AGILE CST-2 Ib.

## METHODS

### Study Design, Sampling and Bioanalytical Methods

Molnupiravir pharmacokinetics were evaluated as part of a phase I dose-escalation study (300, 600, 800 mg twice daily) in patients with PCR-confirmed SARS-CoV-2 infection, within 5 days of symptom onset and presenting with mild or moderate disease (NCT04746183). The study design has previously been described [3] and further details can be found in the Supplementary Material.

Plasma and non-plasma (saliva, nasal secretions and tears) samples were collected at pre-dose, 0.5, 1, 2, and 4 hours post-dose on study days 1 and 5. Plasma samples were collected as previously described [4]. Non-plasma sample collections and molnupiravir dosing conditions are described in the Supplementary Material.

Molnupiravir and NHC concentrations were determined at the University of Liverpool Bioanalytical Facility (Liverpool, UK). Plasma and saliva concentrations were quantified using a validated LC-MS method [4]. NHC in nasal secretions and tears (swabs) were determined using an adaptation of this method described in the Supplementary Material. All assays were validated in accordance with FDA [5] and EMA guidelines [6].

### Pharmacokinetic Data Analysis

Given the small sample size, representation of the pharmacokinetic data was largely exploratory and descriptive. Samples below the lower limit of quantification (LLQ; <2.5 ng/mL) at pre-dose on day 1 were included as 0 ng/mL; those <LLQ beyond pre-dose on day 1 were included as LLQ/2 (1.25 ng/mL). NHC area under the concentration-time curve 0-4 hours (AUC_0-4_), maximum concentration (C_max_) and time to maximum concentration (T_max_) were determined using non-compartmental analysis (Phoenix 64, WinNonlin, v. 8.3, Certara, Princeton, NJ, USA). NHC intercompartmental non-plasma:plasma ratios (R_NP:P_) were calculated on day 1 and 5 for each individual using the plasma as reference (non-plasma AUC_0-4_/plasma AUC_0-4_). Patients without a full profile between 0-4 hours were excluded from AUC_0-4_ summary statistics, and those with sample(s) missing between 0-2 hours were excluded from C_max_ and T_max_ summary statistics.

Linear mixed effects models were applied to evaluate the relationship between log-transformed NHC concentrations in plasma and non-plasma compartments on day 1 and 5 (IBM SPSS Statistics v. 25.0, IBM Corporation, Armonk, NY, USA). Concentrations below assay LLQ were excluded.

## RESULTS

### Patients

Of the twelve participants (n=4 per dosing arm), 9/12 (75%) were female, median (range) baseline age, weight and BMI were 50 years (22-80), 79 kg (54-134) and 29 kg/m^2^ (21-51); time from symptom onset to randomisation and start of treatment was 5 days (3-5).

Ten of the twelve individuals (83%) completed the full treatment schedule. One patient in the 300 mg cohort took 1 of 2 tablets for the second and third dose and a participant in the 800 mg cohort withdrew after the second dose. All pharmacokinetic data were included.

### Non-plasma samples

Molnupiravir was detected at very low concentrations in only 31/106 (29%) plasma and 12/114 (11%) saliva samples [median (range) 5.89 (2.59-27.53 ng/mL) and 4.86 (2.63-31.44 ng/mL, respectively] and therefore not measured in swabs.

In total, 111/113 saliva, 112/112 nasal and 96/106 tear concentrations were included. Sample numbers per cohort are summarised (Supplementary Table 1) and exclusions and samples <LLQ are outlined in the Supplementary Material.

### NHC non-plasma pharmacokinetics

NHC pharmacokinetic parameters are summarised (Table 1). Geometric mean concentration-time profiles are shown (Supplementary Figure 1); additionally, individual profiles are illustrated (Supplementary Figure 2).

**Table 1.** Geometric mean (CV%) NHC pharmacokinetic parameters from plasma, saliva, nasal swabs and tear strips of SARS-CoV-2 infected patients following single (Day 1) and multiple dose (Day 5) molnupiravir 300 mg, 600 mg and 800 mg twice daily (n=4 per dosing arm unless stated otherwise).

| Parameter | 300 mg |  | 600 mg |  | 800 mg |  |
| --- | --- | --- | --- | --- | --- | --- |
|  | Day 1 | Day 5 | Day 1 | Day 5 | Day 1 | Day 5 |
| ***Plasma*** |  |  |  |  |  |  |
| AUC<sub>0-4</sub> (ng.h/mL) | 3031 (45)<sup>#</sup> | 2328<sup>§</sup> | 5690 (22)<sup>*</sup> | 4368 (41) | 8187 (30) | 7005 (21)<sup>*</sup> |
| C<sub>max</sub> (ng/mL) | 1488 (31)<sup>*</sup> | 1048 (17)<sup>#</sup> | 2440 (17) | 1865 (61) | 3447 (32) | 3546 (13)<sup>*</sup> |
| T<sub>max</sub> (h) | 2.00 (1.00-2.00)<sup>*</sup> | 1.00 (1.00-1.00)<sup>#</sup> | 1.00 (1.00-2.00) | 1.00 (1.00-2.00) | 2.00 (2.00-2.00) | 2.00 (2.00-2.00)<sup>#</sup> |
| ***Saliva*** |  |  |  |  |  |  |
| AUC<sub>0-4</sub> (ng.h/mL) | 65 (109)<sup>*</sup> | 106 (93)<sup>*</sup> | 143 (120) | 106 (77)<sup>*</sup> | 289 (52) | 237 (36)<sup>*</sup> |
| C<sub>max</sub> (ng/mL) | 29 (113)<sup>*</sup> | 41 (98) | 73 (127) | 48 (76)<sup>*</sup> | 134 (48) | 109 (27)<sup>*</sup> |
| T<sub>max</sub> (h) | 1.00 (1.00-2.00)<sup>*</sup> | 1.50 (1.00-2.00) | 2.00 (1.00-2.00) | 2.00 (2.00-4.00)<sup>*</sup> | 2.00 (1.00-2.00) | 2.00 (2.00-2.00)<sup>*</sup> |
| R<sub>NP:P</sub> | 0.03 (62)<sup>#</sup> | 0.03<sup>§</sup> | 0.04 (79)<sup>*</sup> | 0.03 (51)<sup>*</sup> | 0.04 (33) | 0.03 (18)<sup>*</sup> |
| ***Nasal Swabs*** |  |  |  |  |  |  |
| AUC<sub>0-4</sub> (ng.h/mL) | 1061 (38)<sup>*</sup> | 673 (27)<sup>*</sup> | 629 (64) | 716 (67) | 2164 (50) | 1611 (73)<sup>*</sup> |
| C<sub>max</sub> (ng/mL) | 805 (70)<sup>*</sup> | 484 (60) | 365 (75) | 321 (65) | 1076 (43) | 738 (87)<sup>*</sup> |
| T<sub>max</sub> (h) | 1.00 (1.00-1.00)<sup>*</sup> | 1.00 (1.00-2.00) | 1.00 (1.00-2.00) | 1.00 (1.00-4.00) | 1.50 (1.00-2.00) | 2.00 (2.00-4.00)<sup>*</sup> |
| R<sub>NP:P</sub> | 0.41 (73)<sup>#</sup> | 0.23<sup>§</sup> | 0.17 (27)<sup>*</sup> | 0.17 (112) | 0.26 (26) | 0.23 (67)<sup>*</sup> |
| ***Tear Strips*** |  |  |  |  |  |  |
| AUC<sub>0-4</sub> (ng.h/mL) | 1731 (44)* | 1071 (38)* | 1137 (96)* | 749 (50)# | 1934 (90)* | 722§ |
| C<sub>max</sub> (ng/mL) | 908 (58)* | 674 (53)* | 411 (100)* | 508 (84) | 985 (95)* | 1267 (40)* |
| T<sub>max</sub> (h) | 0.50 (0.50-2.00)* | 1.00 (0.50-2.00)* | 2.00 (0.50-4.00)* | 1.50 (1.00-2.00)# | 1.00 (1.00-1.00)* | 1.00 (0.50-1.00)* |
| R<sub>NP:P</sub> | 0.77 (36)# | 0.39§ | 0.20 (76)* | 0.17 (47)# | 0.26 (121)* | 0.10§ |

T<sub>max</sub> expressed as median (range)
\* n=3, # n=2, § n=1
AUC<sub>0-4</sub>: area under the concentration-time curve over 0 hours (pre-dose) to 4 hours post-dose; C<sub>max</sub>: maximum concentration; T<sub>max</sub>: time of maximum concentration; R<sub>NP:P</sub>: intercompartmental ratio of non-plasma to plasma (non-plasma AUC<sub>0-4</sub>/plasma AUC<sub>0-</sub>

NHC saliva concentrations were approximately 3% that of plasma [median (range) R_NP:P_ pooled across doses: 0.03 (0.01-0.11); n=16]; the majority of individual ratios were between 0.01-0.04 (n=12). Individual NHC R_NP:P_ for nasal secretions and tears were more variable, (CV%: 60, 70 and 92% for saliva, nasal and tears R_NP:P_. respectively) and overall approximately 6-fold higher than saliva R_NP:P_ [median (range) R_NP:P_ nasal: 0.21 (0.05-0.73); n=17, tears: 0.22 (0.09-1.05); n=12]. Geometric mean (CV%) NHC R_NP:P_, stratified by molnupiravir dose and study day are described (Table 1).

NHC concentrations in saliva and nasal secretions were significantly associated with paired plasma on day 1 and day 5 (*p*<0.0001 for all analyses), whereas statistically significant relationships were not observed for paired tear and plasma NHC concentrations (day 1, *p*=0.068; day 5, *p*=0.344). Time post-dose was included as a repeated effect but addition of random effects for intercept and slope did not improve the models.

## DISCUSSION

Molnupiravir, along with nirmatrelvir/ritonavir (PAXLOVID™), are orally administered antivirals licensed in the UK and with FDA EUA in the US for early treatment of mild-to-moderate COVID-19 in adults with at least one risk factor for developing severe disease. Molnupiravir is currently under phase II evaluation within AGILE including mild-to-moderate COVID-19 without risk factors and in both unvaccinated and vaccinated patients. Molnupiravir is also being investigated for prophylactic use in household contacts of symptomatic COVID-19 patients (MOVe-AHEAD; NCT04939428). Knowledge of drug accumulation within the upper airways and mucosal secretions will inform and support future research in this area.

We observed saliva NHC concentrations that were 3% that of plasma, whereas exposure in nasal secretions and tears was higher at approximately 20% that of plasma (based on pooled AUC_0-4_ ratios). Of the measured saliva, nasal and tear samples, 6, 50 and 61%, respectively were within or above the NHC EC_90_ against SARS-CoV-2 in primary human airway epithelia cultures [7, 8] (approximately 0.5-1 µM ≈ 130-260 ng/mL), suggesting therapeutic concentrations are potentially attained within the nasal and ocular compartments, but not in saliva. However, it is important to note that without established pharmacokinetic/pharmacodynamic relationships or virological data further studies are warranted to determine whether efficacious or prophylactic targets are obtained in non-plasma compartments.

NHC appeared to exhibit a similar absorption and elimination profile in the matrices studied, confirmed by statistically significant linear relationships between plasma NHC with that in non-plasma compartments (with the exception of tears). A strong correlation between saliva and plasma NHC concentrations implies (assuming a one compartment model) that salivary accumulation is dependent upon the plasma concentration. Mucosal permeability and protein binding are major factors in determining salivary drug accumulation, since only unbound drug is available for diffusion into saliva [9]. NHC exists predominantly in unbound form in plasma (unbound fraction ≥0.99) and *in vitro* studies demonstrated that molnupiravir and NHC are not substrates for major drug transporters (e.g. ABCB1; p-glycoprotein) [10, 11]. However, NHC is a substrate for human nucleoside transporters *in vitro* (e.g. CNT1, ENT2) [12] which could modulate non-plasma concentrations of NHC in addition to other factors relating to the drug characteristics or surrounding milieu. Passage of drugs into non-plasma compartments can also be attributed to factors such as pH (e.g. mouth), inflammation (e.g. eye) and flow rate. For example, pharmacokinetics of drug in tears may be affected due to increased lacrimation or infection. Higher turnover or flow rate of saliva may also contribute to the lower concentrations observed. Additionally, the marked variability in nasal and tear NHC concentrations could be associated with the challenging collection procedures.

There are a number of limitations. The small sample size, which is typical of early phase studies, only allowed for a descriptive interpretation of NHC pharmacokinetics and was underpowered for statistical comparisons between matrices. We utilised a truncated sampling schedule between 0-4 hours to limit infection risk, therefore NHC elimination over the 12-hour dosing interval could not be fully assessed in non-plasma compartments. Missing samples led to exclusions from the analysis, particularly for evaluation of R_NP:P_, contributed to data variability and limited data interpretation. Finally, the active triphosphate metabolite, EIDD-2061 was not quantified. Despite the limitations these data add to our understanding of NHC pharmacokinetics, principally at sites of COVID-19 infection.

To our knowledge this is the first report describing penetration of NHC into nasal secretions and tears, and to a lesser extent into saliva. These data support the evaluation of molnupiravir as prophylaxis for SARS-CoV-2 infection.

## Data Availability

The data analysed and presented in the manuscript are available upon reasonable request to the authors

## ACKNOWLEDGEMENTS

The authors wish to thank the patients that participated in the AGILE CST-2 molnupiravir trial and all the staff involved at the NIHR Royal Liverpool and Broadgreen Clinical Research Facility and the team at Liverpool Bioanalytical Facility, who were involved in the processing and analysis of the pharmacokinetic samples, including Elizabeth Challenger and Deirdre Egan. We are grateful to the team at the Southampton Clinical Trials Unit who were involved in study coordination, including Nichola Downs, Geoffrey Saunders, Andrea Corkhill, Kerensa Thorne, Lucy Johnson, Sata Yeats, Kim Mallard, Mike Radford and Keira Fines.

## CONFLICT OF INTEREST

SK has received research funding from ViiV Healthcare, Gilead Sciences, and Merck for the Liverpool HIV Drug Interactions programme and for clinical studies unrelated to the submitted work

GG has received funding from Jannsen-Cilag, Novartis, Astex, Roche, Heartflow, Bristol Myers Squibb, BioNtech; grants and personal fees from AstraZeneca; and personal fees from Celldex, unrelated to the submitted work

WG has received funding from the Wellcome Trust

WH is a cofounder, owner and advisor of/to Ridgeback Biotherapeutics

WP is employed by Ridgeback Biotherapeutics

All other authors have none to declare

