## Supplementary Material for "Pharmacokinetics of ß-d-N4-hydroxycytidine, the parent nucleoside of prodrug molnupiravir, in non-plasma compartments of patients with SARS-CoV-2 infection"

### METHODS

#### *Study Design*

In brief, eligible individuals were randomly assigned to one of three sequential dosing cohorts - molnupiravir dosed orally at 300, 600 and 800 mg twice daily. Within each cohort, patients were randomised at a 2:1 allocation ratio to receive either molnupiravir plus standard-of-care (treatment arms; n=4 per cohort) or standard-of-care alone (control arms; n=2 per cohort). The study was approved by the UK Medicines and Healthcare product Regulatory Agency (MHRA; EudraCT 2020– 001860-27) and West Midlands Edgbaston Research Ethics Committee (20/ WM/0136) and all individuals provided written informed consent. The trial was coordinated by the National Institute for Health Research (NIHR) Southampton Clinical Trials Unit (Southampton UK) with recruitment and treatment of patients taking place at the NIHR Royal Liverpool and Broadgreen Clinical Research Facility (Liverpool, UK).

#### *Pharmacokinetic Sampling*

Patients received their first dose on day 1 and upon return on day 5 in the clinic, patients were instructed to take other doses at home with water. Drug intake was under fasted conditions for a minimum of 2 hours prior to intake and 1 hour after. Medication use was discussed with patients on study visits 3, 5 and 8 and they were asked to bring any unused medication to the visits for pill counts. Patients were also provided with a diary to record drug intake at home which was returned to the clinic for review by the research team.

Saliva samples were collected using Salivette$^{TM}$ tubes (Sarstedt Ltd, UK). The patient was asked to chew on the Salivette swab for approximately 60 seconds after which the swab was returned to the Salivette and centrifuged (within 30 minutes of collection) to yield liquid saliva.

The saliva supernatants (50 µL) were then immediately treated with acetonitrile (acetonitrile:saliva 3:1 v/v) in order to prevent ongoing conversion (via host esterases) of the molnupiravir pro-drug to NHC.

Nasal secretions were collected using 2 x Synthetic Absorptive Matrix (SAM) strips (Mucosal Diagnostics, UK). The applicator was removed from the tube and the absorption end of the swab carefully inserted into the patient's nostril (one each side) with the absorbent strip placed flat against the surface of the inferior turbinate for 60 seconds. The swab was then returned to its original tube and screwed in place using the applicator handle. The weight (to the nearest 0.1 mg) of each SAM strip was determined before and after sampling in order to ascertain the approximate weight of fluid absorbed.

Tears were collected using 2 x Schirmer Tear Test strips which were inserted under the patient's lower eyelid (one in each eye) for 5 minutes. The approximate volume (to the nearest µL) was recorded immediately after collection using the graduated markings on the strip (1-35 µL) and the strip placed inside a clean labelled 2 mL polypropylene tube.

All pharmacokinetic specimens were transported and processed on wet ice and stored at -80°C within 60 minutes of collection.

***Swab Bioanalytical Methods***

In brief, 2 mL of acetonitrile:1mM ammonium acetate (50:50 v/v) was added directly to the tubes containing the swab (calibrators, quality controls and patient samples) and allowed to stand for 1 hour at room temperature. The tubes were sonicated for 30 minutes and exactly 1.8 mL transferred to clean labelled 5 mL glass tubes containing 20 µL of working internal standard solution (2.5 µg/mL $^{13}C_{15}N_2$-N4-hydroxycytidine). Samples were then vortexed, evaporated to dryness under nitrogen at ambient temperature and reconstituted with 200 µL of 1 mM ammonium acetate (adjusted to pH 4.3) ready for injection onto the LC-MS system. Standards and quality controls were freshly prepared on the day of analysis. Working solutions were prepared in 1 x phosphate-buffered saline (PBS) fresh (which served as a surrogate matrix) and 15 µL of each calibrator level pipetted onto a clean SAM or tear strip (in duplicate). The calibration curve ranged between 0.15-75 ng/sample, and was described using a weighted ($1/x^2$) least square linear regression model; recovery was, on average, 96% from nasal and tear swabs and consistent across the calibration range (CV% <15%).

Concentrations of NHC in plasma and saliva were measured in ng/mL, whereas nasal and tear swab NHC were quantified using a ng/sample calibration curve, and converted to ng/mL based on swab volume in µL; a density of 1 was assumed for swab weight to volume conversions.

### RESULTS

#### *Non-plasma samples*

Of the 113 saliva samples, two were excluded due to an incorrect volume of acetonitrile added during processing and ten tear samples of a total of 106 did not have a recorded volume and therefore excluded. All pre-dose concentrations on day 1 (n=12 per matrix) were below assay lower limit of quantification (LLQ; 2.5 ng/mL), as were three 0.5 hour and one 1 hour saliva sample and n=1 tear sample at 0.5 hour post-dose. Additionally, seven pre-dose saliva samples, three nasal swab (n=1 at 0, 2 and 4 hours post-dose) and tear concentrations (n=2 at 0 hours, n=1 at 4 hours) on day 5 were below LLQ.

**Supplementary Table 1** Summary of the number of saliva, nasal and tear samples included in the pharmacokinetic analysis stratified by molnupiravir dosing cohort and study day.

| | **Cohort 1: 300 mg** | | **Cohort 2: 600 mg** | | **Cohort 3: 800 mg** | | **Total** | | |
|---|---|---|---|---|---|---|---|---|---|
| **Samples n/N (%)** | ***Day 1*** | ***Day 5*** | ***Day 1*** | ***Day 5*** | ***Day 1*** | ***Day 5*** | ***Day 1*** | ***Day 5*** | ***Overall*** |
| **Saliva** | 18/19 (95) | 19/19 (100) | 20/20 (100) | 19/20 (95) | 20/20 (100) | 15/15 (100) | 58/59 (98) | 53/54 (98) | 111/113 (98) |
| **Nasal swabs** | 18/18 (100) | 19/19 (100) | 20/20 (100) | 20/20 (100) | 20/20 (100) | 15/15 (100) | 58/58 (100) | 54/54 (100) | 112/112 (100) |
| **Tear strips** | 18/18 (100) | 15/15 (100) | 18/20 (90) | 14/18 (78) | 18/20 (90) | 13/15 (87) | 54/58 (93) | 42/49 (86) | 96/106 (91) |

n: number of samples included; N: total number of samples; %: percentage of samples used in pharmacokinetic analysis

**Supplementary Figure 1**

Geometric mean NHC concentrations over time from saliva (closed points, solid line), nasal swabs (open points, solid line) and tear strips (closed points, broken line) of individuals with SARS-CoV-2 following (**A**) single dose (Day 1) and (**B**) multiple dose (Day 5) molnupiravir 300 mg (circles), 600 mg (squares) and 800 mg (diamonds) twice daily. Data are expressed on a log-linear scale.

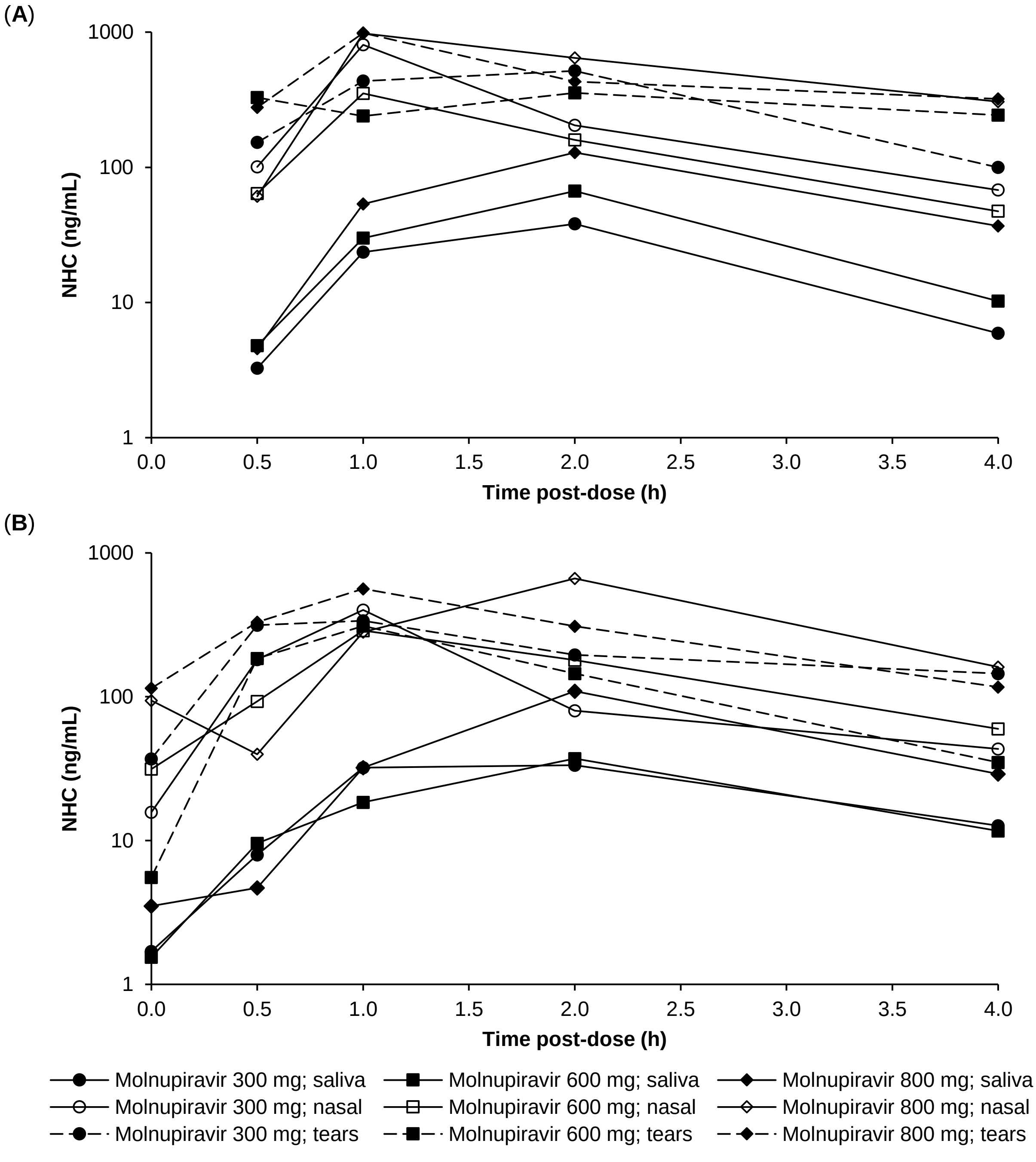

**Supplementary Figure 2** Individual NHC concentration-time profiles from saliva (closed points, solid line), nasal swabs (open points, solid line) and tear strips (closed points, broken lines) of patients with SARS-CoV-2 following single dose (Day 1; left pane) and multiple dose (Day 5; right pane) molnupiravir (**A**) 300 mg, (**B**) 600 mg and (**C**) 800 mg twice daily. Data are expressed on a log-linear scale.

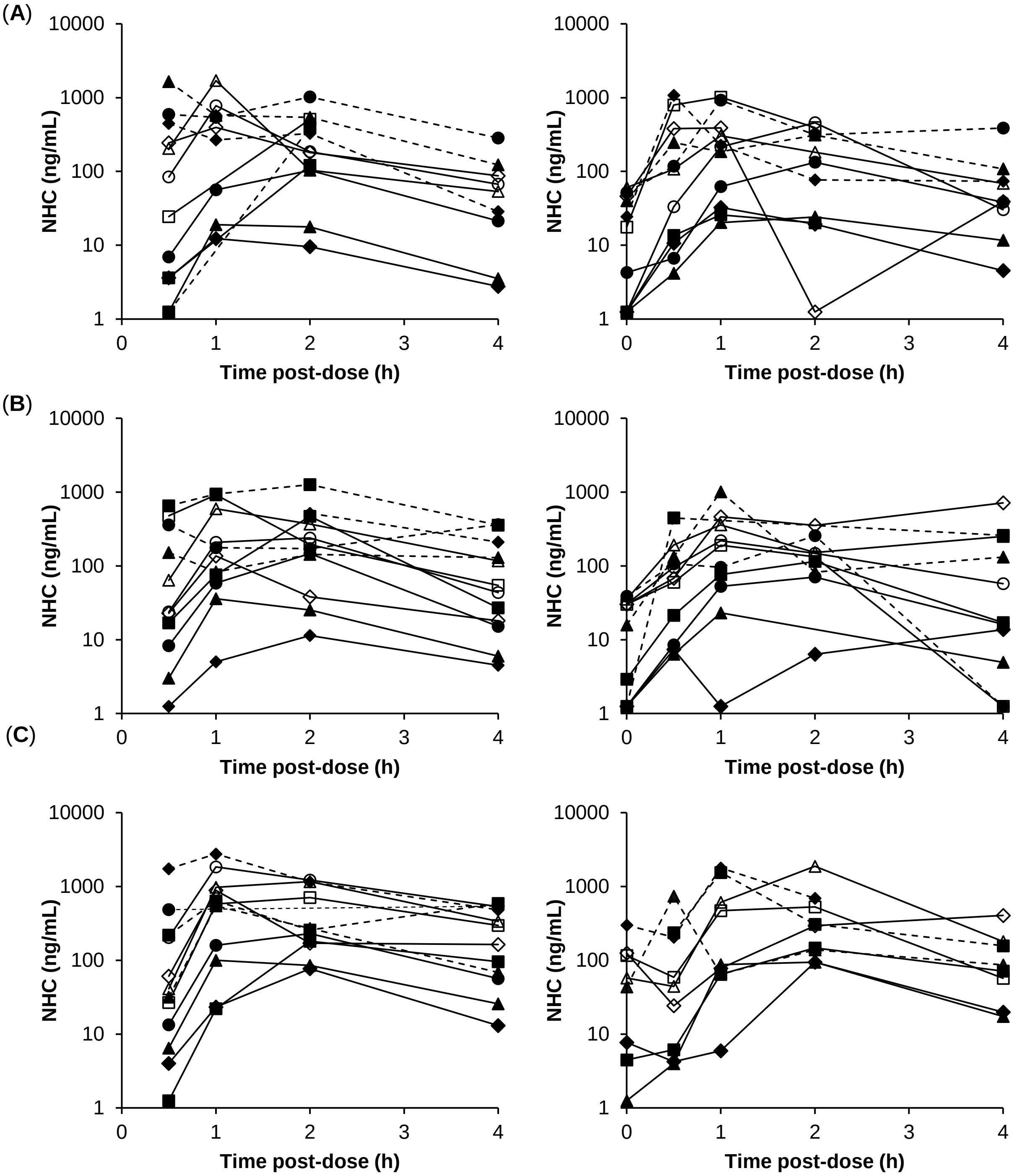
